# Evaluation of the Practicability of a Finger-Stick Whole-Blood SARS-Cov-2 Self-Test Adapted for the General Population

**DOI:** 10.1101/2020.07.13.20152660

**Authors:** Thierry Prazuck, Jean Phan Van, Florence Sinturel, Fredrique Levray, Allan Elie, Denise Camera, Gilles Pialoux

## Abstract

**Background:** COVID-19 (COronaVIrus Disease 2019) is an infectious respiratory disease caused by the novel SARS-CoV-2 virus. Rapid Diagnostic Tests (RDTs) have been developed to detect specific antibodies, IgG and IgM, to SARS-CoV-2 virus in human whole blood and easily usable by the general population are needed in order to alleviate the lockdown that many countries have initiated in response to the growing COVID-19 pandemic. A real-life study has been conducted in order to evaluate the performance of the COVID-PRESTO^®^ RDT and the results have been submitted for publication and are currently under review. Even if this test showed very high sensitivity and specificity in a laboratory setting when used by trained professionals, it needs to be further evaluated for practicability when used by common folk in order to be approved by health authorities for in-home use

**Methods:** 142 participants were recruited between March 2020 and April 2020 among non-medical populations in central France (nuclear plants workers, individuals attending the Orleans University Hospital vaccination clinic and Orleans University Hospital non-medical staff). Instructions for use with or without a tutorial video was made available to the volunteers. Two separate objectives were pursued: evaluation of the capability of participants to obtain an interpretable result, and evaluation of the users’ ability to read the results.

**Results:** 88.4 % of the test users judged the instruction for use leaflet to be clear and understandable. 99.3 % of the users obtained a valid results and according to the supervisors 92.7% of the tests were properly performed by the user. Overall, 95% of the users gave positive feedback toward the COVID PRESTO^®^ as a potential self-test. No influence of age and education was observed.

**Conclusion:** COVID-PRESTO^®^ was successfully used by an overwhelming majority of participants and its utilization was judged very satisfactory, therefore showing a promising potential as a self-test to be used by the general population. This RDT can become an easy-to-use tool to help know whether individuals are protected or not, particularly in the perspective of a second wave or a mass vaccination program.

## Introduction

In Wuhan, China the end of the year 2019 marked the emergence of a new type of pneumonia caused by a then-unknown agent. Shortly after the first reports of the disease, the Chinese health authorities and the World Health Organization announced that a newly-discovered type of coronavirus was responsible for the disease. This new virus was named SARS-CoV-2 (Sever Acute Respiratory Syndrome-CoronaVirus-2). This virus is a new member of the coronavirus family that already includes SARS-CoV and MERS-Cov responsible for the SARS outbreak in 2003, and an ongoing outbreak that started in 2012 in the Middle East, respectively.

The disease caused by the SARS-CoV-2 virus is called COVID-19 (COronaVIrus Disease-2019). The site of infection is located on the upper/lower respiratory tract [1]. The mean incubation period is approximatively 5.2 days and the most common symptoms are dry cough, fever and fatigue. Other symptoms include anosmia (loss of smell), ageusia (loss of taste), headache, sore throat and in the most severe cases acute respiratory distress syndrome.

Within the last 6 months, according to the Johns Hopkins University Coronavirus resource center the COVID-19 pandemics has spread over 188 countries; leaving very few regions of the world untouched. At the end of January 2020, the WHO has declared this outbreak a global health emergency and the mark of 100 000 deaths was reached on April 12^th^ leading to a little less than 350 000 deaths on May 26^th^ for a total of 5 519 878 confirmed cases. Governments have taken extreme measures to try slow the spreading of the virus by imposing strict social distancing rules and it is estimated that 1.7 billion people have been confined home worldwide.

In most countries, these measures have been effective in slowing down the transmission of the virus. In May 2020, several governments started easing the confinement rules but questions remain in regards to how to test, control and track every case in a large population at least until an effective treatment or vaccine can be found. To this end, the development of new testing tools that could be distributed at a large scale in the population is crucial. Serological tests able to detect the specific antibodies against the SARS-CoV-2 in people seem to be particularly good candidates because there are fairly easy to use and execute without an extensive training.

The AAZ COVID-PRESTO^®^ is a Rapid Diagnostic Tests (RDTs), easy-to-use device based on lateral flow chromatographic immunoassay from a single drop of blood (Figure 1). It has been evaluated in terms of sensitivity and specificity and showed good results that have been submitted for publication and currently under review [2]. Those results were obtained in a controlled setting where the test was performed by trained professionals. In order to be suitable as a self-test for the general population, COVID-PRESTO^®^ has to be evaluated for practicability, i.e. its capacity of being able to be performed by untrained individuals. The aim of this study was to evaluate the COVID-PRESTO® test in terms of participants’ capability to obtain an interpretable result, and the users’ ability to interpret the results.

**Fig 1.**
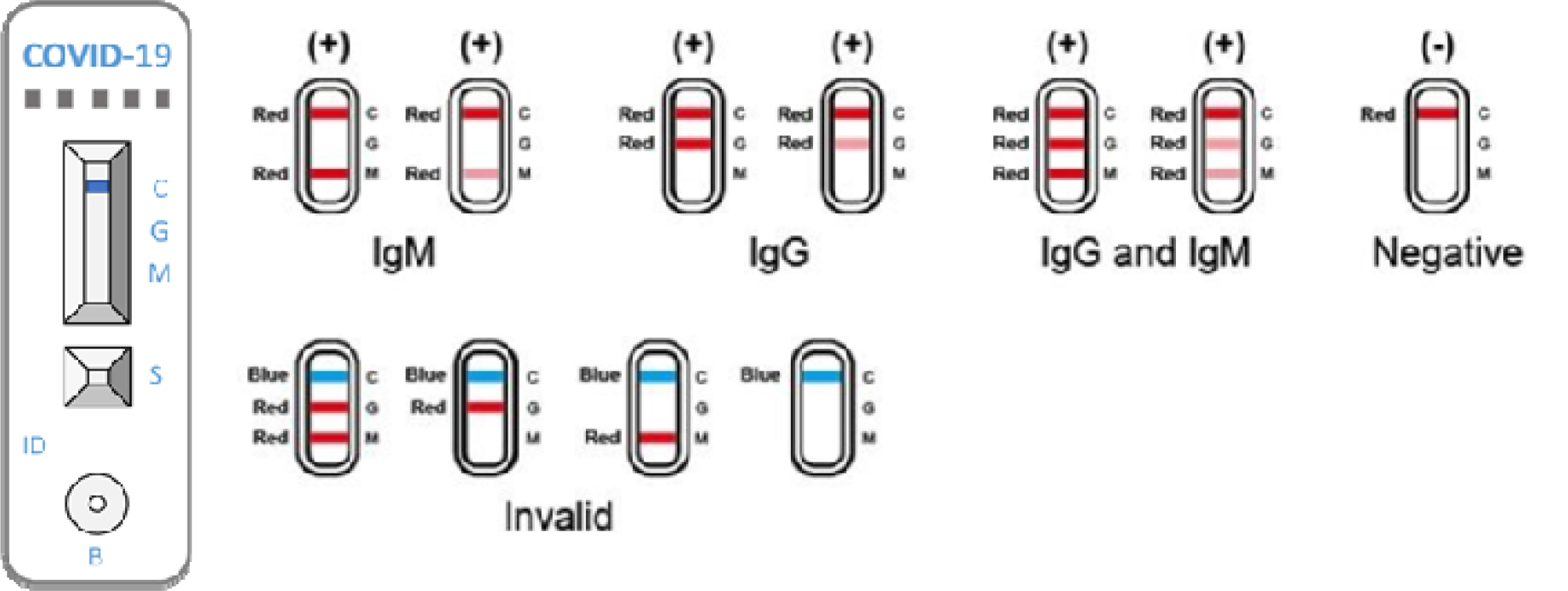
Interpretation of results for COVID-PRESTO^®^.

## Methods and Materials

### Ethics Approval

The study was approved by the Regional Orléans Research and ethics Committee on March 17^th^ 2020, and informed consent was obtained from each participant.

### Study population

The study volunteers were selected from four different locations in Central France from March 20th 2020 to May 5th, 2020: two nuclear plants, individuals visiting the vaccination clinic of Orleans Regional Hospital and non-medical staff from Orleans Regional Hospital. The decision of recruiting non-medical staff exclusively was made in order to avoid bias due to previous or current experience in blood drawing. A total of 142 volunteers participated to the study. Two were excluded from the analysis because they did not fill in the questionnaire.

A 2 steps design similar to the design used to validate an HIV self-test was used for this study [3].

### Substudy 1: Usage of the self-test

Each volunteer was invited to read the test’s information for use replaced and/or associated to an instructional video in order to give clearer instructions.

The instructions were the following:

Every volunteer had to use the lancet needle to prick the side of the fingertip to let a large drop of suspended blood form, collect the drop with a 10 µl capillary micropipette that filled automatically, transfer the blood into the sample well and finally, add two drops of buffer in the buffer well.

Each participant was asked to fill in a satisfaction questionnaire on the test as a future self-test intended for the general population (Table 2).

**Table 1:**
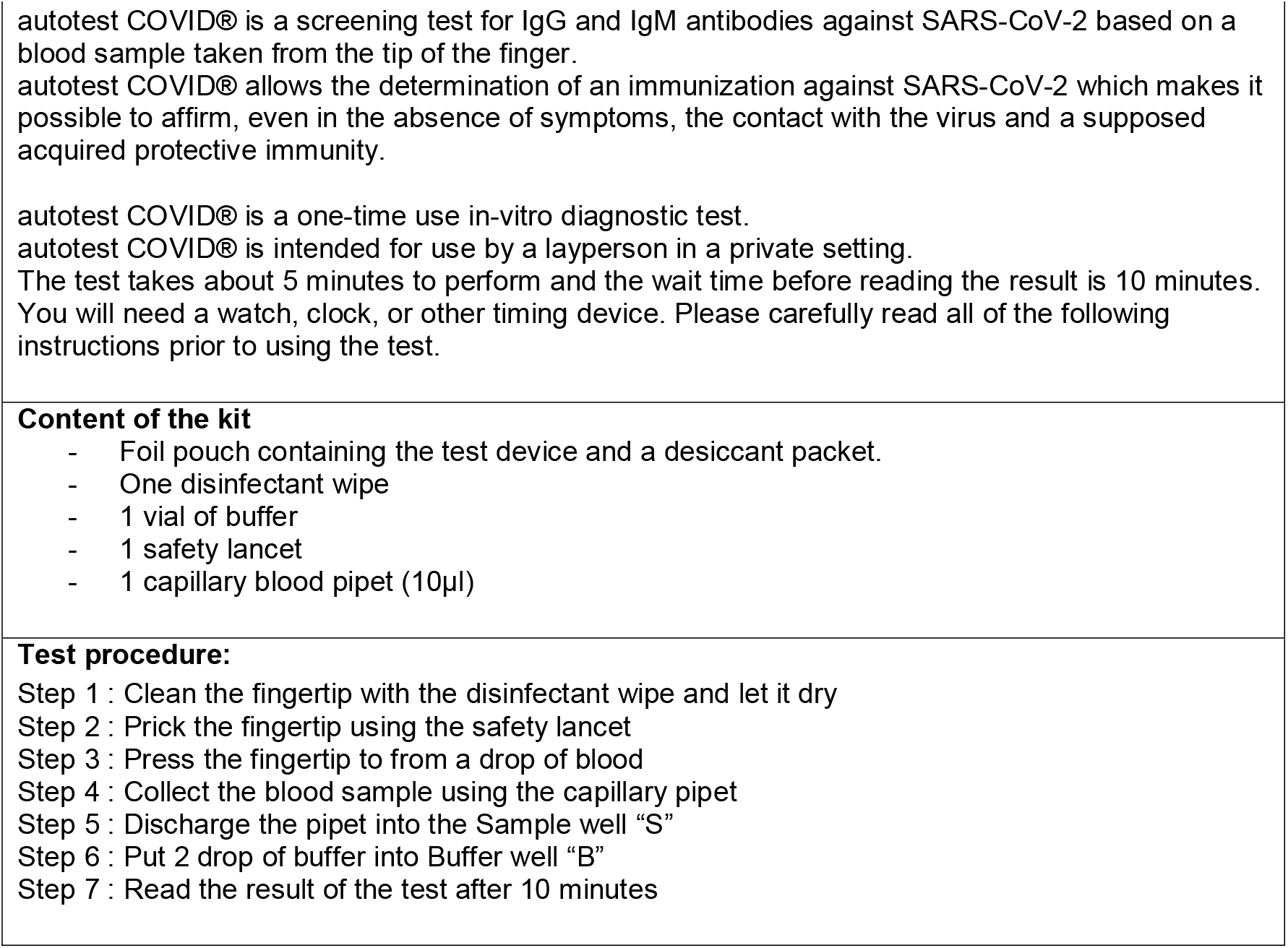
Instruction for use.

|  |
| --- |
| **Table 1: Instruction for use.** |
| autotest COVID® is a screening test for IgG and IgM antibodies against SARS-CoV-2 based on a blood sample taken from the tip of the finger.<br>autotest COVID® allows the determination of an immunization against SARS-CoV-2 which makes it possible to affirm, even in the absence of symptoms, the contact with the virus and a supposed acquired protective immunity. |
| autotest COVID® is a one-time use in-vitro diagnostic test.<br>autotest COVID® is intended for use by a layperson in a private setting.<br>The test takes about 5 minutes to perform and the wait time before reading the result is 10 minutes.<br>You will need a watch, clock, or other timing device. Please carefully read all of the following instructions prior to using the test. |
| **Content of the kit** |
| - Foil pouch containing the test device and a desiccant packet.<br>- One disinfectant wipe<br>- 1 vial of buffer<br>- 1 safety lancet<br>- 1 capillary blood pipet (10µl) |
| **Test procedure:** |
| Step 1 : Clean the fingertip with the disinfectant wipe and let it dry<br>Step 2 : Prick the fingertip using the safety lancet<br>Step 3 : Press the fingertip to form a drop of blood<br>Step 4 : Collect the blood sample using the capillary pipet<br>Step 5 : Discharge the pipet into the Sample well "S"<br>Step 6 : Put 2 drop of buffer into Buffer well "B"<br>Step 7 : Read the result of the test after 10 minutes |

**Table 2.** Satisfaction questionnaire.

| Satisfaction Questionnaire |
| --- |
| **1.** Did you find the test's instructions for use that was provided to you, clear and comprehensible? |
| ☐ Yes ☐ No |
| **2.** Did you find the instructional video, clear and comprehensible? |
| ☐ Yes ☐ No |
| **3.** Did you manage to collect a sufficient drop(s) of blood? |
| ☐ Yes ☐ No |
| **4.** Did you manage to fill the pipette? |
| ☐ Yes ☐ No |
| **5.** Did you manage to deposit the blood and buffer in the wells without mistake? |
| ☐ Yes ☐ No |
| **6.** Did you obtain a result (at least one band)? |
| ☐ Yes ☐ No |
| **7.** What is your opinion regarding the utilization of this test as a self-test? |
| ☐ Bad ☐ Mediocre ☐ Good ☐ Very good ☐ Excellent |
| **8.** What is your opinion regarding the readability and the interpretation of this test? |
| ☐ Bad ☐ Mediocre ☐ Good ☐ Very good ☐ Excellent |

The test user was monitored by an observer (Occupational health physician in the nuclear plants, trained nurse or physician in the Orleans Regional hospital) who could assist the user, if asked to, and give his/her feedback on the execution of the different tasks. This feedback could be different from the user’s personal opinion (Table 3).

**Table 3.** Supervisor’s Opinion.

| Supervisor's Opinion |
| --- |
| **1.** Was the supervisor's assistance requested by the participant? |
| ☐ Yes ☐ No |
| **2.** What is your opinion regarding the execution of this test by the participant? |
| ☐ Bad ☐ Mediocre ☐ Good ☐ Very good ☐ Excellent |

### Substudy 2: Reading and interpretation of the results

Each participant was shown a basket containing 6 standardized test results (2 positives, 2 negatives and 2 invalids). The participant had to randomly choose three out of the six standardized tests and write down the results for each test. A supervisor was in charge to collect the responses and assess their correctness.

### Data Analysis

Population were described in terms of %, and mean values standard deviation, range and median values. Demographics characteristics such as age and education level, associated with the responses to the questionnaire were evaluated using univariate and stratified analyses and the Cochran-Mantel-Haenszel test. In case of overall differences between groups, *post hoc* Fisher or χ^2^ tests with Bonferroni correction were applied for two-group comparisons.

## RESULTS

Overall, 142 volunteers were selected to participate to the study, among those 2 did not fill in the Satisfaction questionnaire and 96 participated to Substudy 2. The distribution of gender was in favor of men, twice as much represented than women. Four different age classes were defined 20-29, 30-39, 40-49 and ≥50. Three education level classes were defined according the toe International Classification of Education (2011 version): Level 3 or below, Level 4 or 5, and Level or above.

The demographics characteristics are provided in Table 4:

**Table 4.** Demographics characteristics of the study population.

| Characteristics | Substudy 1 |  | Substudy 2 |  |
| --- | --- | --- | --- | --- |
|  | N=142 |  | N=96 |  |
|  | N (%) | % | N (%) | % |
| **Study Site:** |  |  |  |  |
| Dampierre Nuclear Plant | 45 | 32.1 | 23 | 24.0 |
| St Laurent Nuclear Plant | 37 | 26.4 | 15 | 15.6 |
| Orleans Regional Hospital | 58 | 41.4 | 58 | 60.4 |
| **Age** |  |  |  |  |
| 20-29 | 32 | 22.5 | 25 | 26.0 |
| 30-39 | 45 | 31.7 | 30 | 31.3 |
| 40-49 | 46 | 32.4 | 29 | 30.2 |
| $\geq 50$ | 19 | 13.4 | 12 | 12.5 |
| **Gender** |  |  |  |  |
| Male | 95 | 67.8 | 56 | 58.3 |
| Female | 45 | 32.2 | 40 | 41.7 |
| **Education Level\*** |  |  |  |  |
| Level 3 or below | 23 | 16.4 | 18 | 18.8 |
| Level 4 or 5 | 75 | 53.6 | 48 | 50.0 |
| Level 6 or above | 42 | 30.0 | 30 | 31.3 |

\*According to the International Standard Classification of Education 2011

### Substudy 1

A total of 103 participants read the instructions for use provided with the test. Among those 67 volunteers read the instructions and watched the video. 91 participants (88.4%) found the written instructions to be clear and comprehensible. 106 participants watched the video making it 39 volunteers that watched the video as the only instruction medium. Among these 106 volunteers, 96 (90.6%) found the video to be clear and comprehensible. A subgroup analysis showed that there was no difference on the comprehension of the instructions notice when age or education level was considered. However, there was a statistical difference in the comprehension of the instructional video among the groups. Indeed, the video was judged comprehensible by only 77.8% of the participants aged 29 or less compared to 91.7% (30-39), 96.7% (40-49) and 100.0% (≥50) by the other groups (Cochran-Mantel-Haenszel test’s overall p= 0.0465). However, *post hoc* Fisher’s tests with Bonferroni correction revealed that the proportion of users that found the video comprehensible in the 20-29 range was not significantly lower when groups were compared by pairs, meaning that age has little influence on the comprehension of the instructional video. There was also an overall statistical difference in the comprehension of the video when education level was considered as an influencing factor (Cochran-Mantel-Haenszel test’s overall p= 0.0235). Nevertheless, when two-group comparisons were performed using a Fisher’s t test or a Chi^2^ with a Bonferroni correction, no difference was observed. These results show that education level did not have an effect on the understanding of the instructional video. Taken together these results show that the instructions were well received and understood by all participants with little to no impact of age or education level on their comprehension.

The COVID-PRESTO^®^ technical practicability was assessed by analyzing the participants’ responses to the satisfaction questionnaire. 130 out of the 140 participants (92.8%) who filled in the questionnaire, were able to draw enough blood using the lancet needle on their fingertips. However, the ten participants that could not collect the recommended quantity of blood were able to proceed with the subsequent steps. 128 participants (91.4%) succeeded in filling up the pipette with blood on the first try and without assistance. 135 testers (96.4%) were able to correctly deposit both the blood and the buffer in their respective wells. Among those, 7 participants needed assistance to fill the pipette. At the end of the procedure, nearly all participants (139/140, 99.3%) declared to have been able to get a valid result (defined by a pink band in the control lane).

In terms of overall satisfaction, 133/140 participants (95%) rated positively the COVID- PRESTO^®^ (defined as “Good”, “Very Good” or “Excellent”), based on the ease of execution of the test’s procedures. 45% of the participants rated the test as good and 50% as “Very Good” or “Excellent”. A subgroup analysis revealed that age and education had no impact whatsoever on the overall satisfaction towards the COVID-PRESTO^®^ test.

Supervisors were also asked to answer a survey after each participant was done with the test’s procedures. It is however, important to point out that only 122 answers to the supervision survey were collected. During these supervised sessions, 92/122 (76.4%) participants did not ask for assistance to the supervisor. Only 30 participants requested an assistance mainly for tips on how to take off the cap of the needle as well as on how to use the pipette. The subgroup analysis showed that almost half of the participants (43.8%) with an education level of 4 or 5 requested assistance and this proportion was statistically higher when compared to the proportion of participants having requested assistance in the other groups (Overall Cochran-Mantel-Haenszel test’s p = 0.0004; χ^2^ p value of 0.009 and 0.006 *vs*. level ≤ 3 and ≥ 6 respectively).The supervisors’ opinion on the execution of the procedures by the participants was collected and the execution was rated as good or better for 113/122 (92.7%) volunteers. 45.9% of the volunteers were rated as “Good”, 36.1% as “Very Good” and 10.7% as “Excellent”. When age and education parameters were considered during a subgroup analysis, no difference were observed between the categories in terms execution ratings.

### Substudy 2

96 volunteers took part of this second phase down from the 140 participants in the first phase. The volunteers had to randomly choose 3 tests out of 6 and assess their readability. Among these 96 participants 94 (97.9%) judged the readability of the sorted test to be good or better (“very good” or “excellent”). 41.2% of the volunteers rated the test legibility as “Good”, 41.7% as “Very Good” and 14.5% as “Excellent”. Only two individuals incorrectly interpreted the test by judging an invalid test as valid. The confusion originated from the fact that they had not understood that the control lane had to be pink instead of blue. Overall, 288 tests were read and only 2 were not interpreted correctly resulting in a 99.3% success rate. These two participants were 26 and 42 years old and with an educational level of 3 and 4 respectively. A subgroup analysis considering the age and education level classes showed that these two parameters don’t have any influence of the ability of the participant to correctly interpret the test results.

## DISCUSSION

This study had the objective to test the adequacy of one Rapid Diagnosis Test for Covid-19 in regards to a potential release for the general population. This test was developed to detect the presence of antibodies targeted against the SARS-CoV-2 as an indirect marker of prior infection.

The use of such tests with the help of appropriate instructions and interpretation guidelines could prove to be essential in supporting the current public health effort. Indeed, these tests would provide intelligence data on the dissemination of virus across the population and therefore would a valuable tool to help comprehend the epidemics. The use of such tests will be also essential to know immune status of patients in view of a second wave or a possible mass vaccination program [4].

An extensive list of the test available worldwide can be found in the FIND foundation website (https://www.finddx.org/covid-19/pipeline/?section=immunoassays#diag_tab). Around a hundred of COVID-19 serological tests are currently commercially available in the US through the Emergency Use Authorization (EUA) granted to the US Centers for Disease Control and Prevention by the Food and Drug Administration (FDA) [5]. The FDA has provided guidance for manufacturers of serological test in order to promote and facilitate rapid market access on the basis that such tests could help provide crucial information about the prevalence of COVID-19 infections in different communities [6]. However, an NIH independent evaluation has shown that a concerning number of commercial serological test are not being appropriately promoted or show poor performance [6]. In Europe, according to regulations, manufacturers should submit data regarding “handling suitability of the device in view of its intended purpose for self-testing” for assessment by notified bodies before making it available to the public [7]. The European Centre for Disease Prevention and Control reports that several COVID-19 RDT are being marketed with incomplete and event sometimes fraudulent documentation and unsubstantiated claims, some of these test being sold as self-tests and thus, several European countries has banned the marketing of such tests until further notice [8] Therefore, there is a crucial need of more documented studies regarding those tests [9,10].

From a regulatory standpoint, “self-testing” is a more stringent regulated category of *in vitro* diagnosis tests compared to the regular kind, intended for professional use only, and the French health authority (Haute Autorité de Santé, HAS) estimates that in absence of reliable data regarding the available self-tests it is premature to promote their use [11]. COVID- PRESTO^®^ aims to be one of the first SARS-CoV-2 serological test to be officially approved as a reliable and accurate self-test in order to offer a viable option for large scale testing. It is important to keep in mind that these tests are not designed to detect an ongoing infection but rather a prior infection which would give intelligence about the true prevalence of the virus. However, a positive test will not mean that the tested individual is no longer infectious but will give the information that at some point in the past. As of today, there is no solid evidence that a prior infection will offer a strong immunity towards a second exposure and if so for how long [12], nevertheless, a pre-published study demonstrated that most of the patients having presented a mild form of the COVID-19, developed neutralizing antibodies [4].

COVID-PRESTO^®^ has recently been evaluated for performance and showed a sensitivity ranging from 69% for patients with symptoms that occurred from 11 to 15 days before the date of test and 100% in patients who experienced first symptoms more than 15 days before the test. These results are currently being reviewed for publication [2]. However, for it to be approved as a self-test, a practicability study was needed in order to assess the feasibility of the test by untrained individuals with different education levels. First of all this study showed that the instruction materials provided with the test are clear and comprehensible regardless of the user’s age or education level. However, our results showed that age or education level may slightly influence the comprehension of non-written instructions, i.e. instructional video. This finding indicates that it may be necessary to include a variety of instruction media along with the test to ensure its comprehension by a broader audience. These media may include written instructions, cartoons or videos.

We observed that the education level influenced the fact that whether or not the participant asked for assistance. Nevertheless, no influence of the age nor the education level on either the ability to correctly read the test or to execute the procedures according to the supervisors, was observed in our study. This strongly suggest that the execution of the test is accessible to a wide range of persons. Finally, this study revealed that the COVID-PRESTO^®^ is judged practical with a global satisfaction rate of 95% by the users and is favorably seen as a potential self-test.

Data are scarce regarding the performance of the existing serological COVID-19 test and to our knowledge this is the first practicability study regarding a COVID-19 RDT making it difficult to compare our results to other devices. However, that this kind of study is necessary before making a self-test available to general public in order to, in one hand, avoid confusion about false positive results thus leading to unnecessary demands of health services and, in the other hand, avoid false negative results potentially leading to underestimation of the virus presence across population.

The ease of understanding the instructions is always a challenge when designing a self-test but our study shows that the COVID-PRESTO^®^ test procedures fare well in that regard. However, feedback from users showed that there is still room for improvement regarding the instructions and video. Indeed, some of the volunteers had legitimate questions on technical procedures such as the handling of the lancet needle, and the use of the pipette. It is important to point out that in our study, socio-demographic parameters such as age and education, did not influence any of the tested parameters suggesting that the instructions as well as the procedures are sufficiently clear and simple to be executed by the general public. The next step for a person that has been tested positive for the Covid-19 is knowing if he/she is still contagious and whether he/she has to isolate from his/herself from others. Given the data currently available regarding the SARS-CoV-2, although it has been recently demonstrated that individuals who have recovered form a mild form of the COVID-19 possess neutralizing antibodies [4], we can only assume without certainty that infection with the virus generates protective immunity [12],. It is therefore important that the test instructions includes clear and understandable guidance regarding the actions that need to be taken in case of a positive or negative results, in order to avoid relaxation of the safety measures. To this aim, in case of a positive IgM test, the instructions for use of the COVID- PRESTO^®^ test will recommend to seek care from a Primary Care provider and undergo further testing (PCR) to confirm/invalidate the presence of an active infection. In case of a confirmed active infection, the instructions will enjoin the patient to observer a strict 14 days quarantine.

## Conclusion

These 2 substudies indicate that the finger-stick COVID PRESTO^®^ self-test is practical and that test users correctly read the results. The COVID-PRESTO^®^ should be considered as a suitable candidate for a public release in order to provide an additional tool to gather information about the dissemination of the virus across the population.

## Data Availability

data will be available without restriction at clinact.com, a french CRO

## Contributions

Conceived and designed the experiments: TP and GP. Recruited the participants and conducted the study: JPV, FS, FL, AE and DC. Analyzed the data: TP and GP. Wrote the manuscript: TP and GP.

## Acknowledgments

The authors would like to thank the technical staff of the Department of Infectious diseases for excellent assistance. Furthermore, the authors thank Thibaut de Sablet of Clinact, France for providing medical writing support/editorial support in accordance with Good Publication Practice (GPP3) guidelines.

